# Trends in childhood vaccination coverage in the European Union, 1980–2024: between long-term increases and recent decreases

**DOI:** 10.1101/2024.11.13.24317259

**Authors:** Leonardo Villani, Luigi Russo, Francesco Andrea Causio, Cosimo Savoia, Roberta Pastorino, Chiara De Waure, Walter Ricciardi, Stefania Boccia

**Affiliations:** Section of Hygiene, University Department of Life Sciences and Public Health – Università Cattolica del Sacro Cuore, Rome, Italy; UniCamillus - Saint Camillus International University of Health and Medical Sciences; Department of Woman and Child Health and Public Health, A. Gemelli University Hospital Foundation, IRCCS, Rome, Italy; Department of Medicine and Surgery, University of Perugia, Perugia, Italy

**Author notes:** **Corresponding Author:** Luigi Russo, MD, Section of Hygiene, University Department of Life Sciences and Public Health - Università Cattolica del Sacro Cuore. Largo Francesco Vito 1, 00168, Rome, Italy.

**Keywords:** Vaccination, childhood, European Union, public health, trend analysis

## Abstract

**Background:** Vaccination is among the most effective public health interventions, yet vaccine hesitancy in the European Union (EU) has driven declining coverage and resurgences of vaccine-preventable diseases. Trend assessments are essential for informing strategies to maintain high coverage levels.

**Methods:** We analysed WHO/UNICEF Estimates of National Immunization Coverage data (as of September 2025) for 27 EU countries from 1980 to 2024. Coverage for seven first-year antigens (DTP-3, HEPB-3, HIB-3, POL-3, PCV-3, MCV-1, RCV-1) was evaluated using joinpoint regression for Average Annual Percent Change (AAPC) overall and recent Annual Percent Change (APC) shifts.

**Findings:** Of 183 country-vaccine trends, 128 showed positive AAPCs but recent APCs indicated predominant declines (110 negative evolutions). In 2024, only 71 combinations reached ≥95% coverage; Luxembourg achieved this for all antigens. Seventeen countries (e.g., Germany, Romania) had declines in ≥4 vaccines, especially HIB-3, POL-3, and DTP-3.

**Interpretation:** Despite long-term gains, recent EU-wide coverage declines signal urgent risks to herd immunity. National and community actions are needed to reverse trends.

**Funding:** None.

## Introduction

Vaccinations represent one of the most successful and cost-effective public health interventions and have led to the eradication, elimination, and control of many infectious diseases in the last few decades (1,2). The relevance of vaccination is testified by its contribution to eradicating smallpox in 1980, while nowadays it is a key tool for eliminating wild polio (3,4). Vaccinations have reduced the burden of diseases such as tetanus, whooping cough, pneumonia, measles, influenza, and diphtheria, annually preventing 4-5 million deaths worldwide (5), and over 154 million deaths, especially among children, in the last 50 years vaccinations (6). In addition, vaccination is one of the most cost-effective interventions, reducing direct and indirect costs related to infectious diseases (7,8).

In recent years, however, vaccine hesitancy, defined as a delay in acceptance or outright refusal despite the availability of vaccination services (9), has gained momentum, especially due to misinformation at both political and scientific levels. This phenomenon affects countries worldwide differently and unevenly, and depends on several social, political, educational, economic, behavioural, and health factors (9). In this context, the increase of hesitancy, as well as the disruption caused by the COVID-19 pandemic (10), has led to a reduction in vaccine coverage, with vaccine-preventable diseases re-emerging, even in high-income European Union (EU) countries (11). To face this situation, many countries adopted several measures, with high variance within EU countries, with some recommending some vaccines and some adopting vaccination mandates (12). In particular, 13 EU countries (Belgium, Bulgaria, Croatia, Czechia, France, Germany, Hungary, Italy, Latvia, Malta, Poland, Slovakia and Slovenia) have established at least one mandatory vaccination for school entry, with differences in terms of modalities, timing, and the number of vaccines (12). For example, while in Belgium, the only mandatory vaccination is against polio, in place since 1967, in Italy the number of mandatory vaccinations increased in 2017 from four to ten (13), and in France, a law of 2018 established eleven mandatory vaccinations (14). Other countries have maintained an approach emphasizing personal freedom, aiming at empowering citizens and encouraging them to take the best decision. In Sweden, after experiencing limited outbreaks of measles and rubella in 2013 (15), new vaccine policies were introduced, offering immunization programmes to groups at-risk and investigating underserved populations with low vaccination coverages.

Currently, information on population coverage against several preventable communicable diseases at country level is available from different sources, such as international organisations, Ministries of Health, national institutions, and independent research centres. However, to date, a systematic synthesis of EU vaccination coverage trends has not been developed. Nevertheless, analysis of this data is crucial to controlling the evolution of infectious diseases and to assessing the effectiveness of adopted targeted policies at the national and international levels (16–18). Evaluating trends in vaccination coverage allows for the provision of scientific evidence for policymakers to implement effective and targeted prevention strategies.

In this context, our study aims to describe the trend of vaccination coverage for seven vaccinations from 1980 to 2024 across EU countries, and to identify positive, negative, or stationary evolutions for both the single vaccination and the single country.

## Methods

### Data source

We extracted data about vaccination coverage from the publicly available WHO/UNICEF Estimates of National Immunization Coverage (WUENIC) dataset, based on data officially reported to the organizations by Member States every year, as of September 2025 (19–21), and we included all 27 EU countries in our analysis. We classified the 27-EU countries with the classification of EuroVoc, as shown in Supplementary table 1. We considered the period from the first year available, 1980 (or more recently if data were unavailable) to 2024 (or the most updated data when no data were available for 2024). Based on the availability of data, we considered coverage indicators referring to seven vaccinations scheduled during the first year of life to prevent nine vaccine-preventable diseases. In particular, we selected the vaccination for diphtheria, tetanus toxoid, and pertussis (DTP-3), hepatitis B (HEPB-3), Haemophilus influenzae type B (HIB-3), measles (MCV-1), pneumococcus (PCV-3), polio (POL-3), and rubella. Further description of the vaccination schedule, according to the WHO (22), is available in Supplementary table 2.

### Statistical analysis

We conducted descriptive statistics and preliminary comparisons across vaccination coverage during the considered in each country. To assess changes in vaccination coverage trends across EU countries, we used the Joinpoint regression model. Joinpoint regression is a tool for analysing changes in trends over time and, for this reason, it is used in many scientific domains (23–25), including vaccination (26–28). Indeed, this model allows for identifying a point (“joinpoint”) when a change in a parameter occurs within the time series. In our study, we conducted a trend analysis for each coverage indicator for all countries. In particular, we considered the year as the independent variable and the vaccination coverage indicator as the dependent variable, expressed as a percentage, while the results were sorted by country. Moreover, the joinpoint model allows for estimating an Annual Percent Change (APC) in vaccination coverage, reflecting an increase or decrease over time. For each vaccination, the presence of any joinpoint (the software ranges from 0 to a maximum of 7) expressed changes in the APC trend. Finally, the model estimates the Average Annual Percent Change (AAPC), a summary measure of the trend over the entire period. We considered the APC and AAPC significant when p < 0.05. The analysis was performed using Joinpoint Trend Analysis Software 5.3.0 – Desktop Version, available from the Surveillance Research Program of the US National Cancer Institute (29).

### APC and AAPC analysis

The last two trends of the APC that occurred in the time series were considered for each coverage indicator. For reasons of consistency, we called the second-last trend “trend A”, accounting for the period between the second-last and the last joinpoint, and the last trend “trend B”, accounting for the period after the last joinpoint. Trends A and B are marked for clarity in Supplementary Figure 1.

Based on the significance of the APC, each trend was labelled as follows compared with vaccination coverage: significant increase, significant decrease, non-significant increase, and non-significant decrease. Variation between trends A and B was considered when defining the confirmation or reversal of the trend for each coverage indicator in each country.

For each indicator, based on the changes between trend A and trend B identified through joinpoint analysis, two possible evolutions were considered: positive or negative. In detail, the trend was classified as positive when trend A remained significantly positive in trend B, when trend A of any type became significantly positive, or when a significantly negative trend A lost its significance in trend B. Conversely, a pair of trends was classified as negative when trend A remained significantly negative in trend B, when trend A of any type became significantly negative, or when a significantly positive trend A lost its significance in trend B. If the change between trends A and B was not significant, the pair was defined as mildly positive when trend A remained non-significantly positive in trend B or when a non-significant negative trend A became non-significant positive in trend B, and as mildly negative when trend A remained non-significantly negative in trend B or when a non-significant positive trend A became non-significant negative in trend B. For trends without a joinpoint, the trend was classified as positive (significant increase), mildly positive (non-significant increase), negative (significant decrease), or mildly negative (non-significant decrease).

Finally, pairs of trends classified as positive or mildly positive were considered increasing, while those classified as negative or mildly negative were considered decreasing. Eventually, to identify which EU countries and which antigen presented an overall improving or worsening trend in vaccination coverages, we considered the AAPC for each indicator, identifying both positive and negative evolution.

## Results

### Overall results

Out of 189 possible vaccination–country trends (7 vaccinations across 27 EU countries), 183 analyses were performed (Supplementary Figures 2-5); data were unavailable in 6 cases. Overall, 55 trends showed a negative AAPC and 128 a positive AAPC, indicating a general increase in coverage from the first year of available data. Among these, 43 negative AAPC were statistically significant, whereas 110 positive AAPC were significant. All AAPCs are shown in Table 1.

**Table 1.**
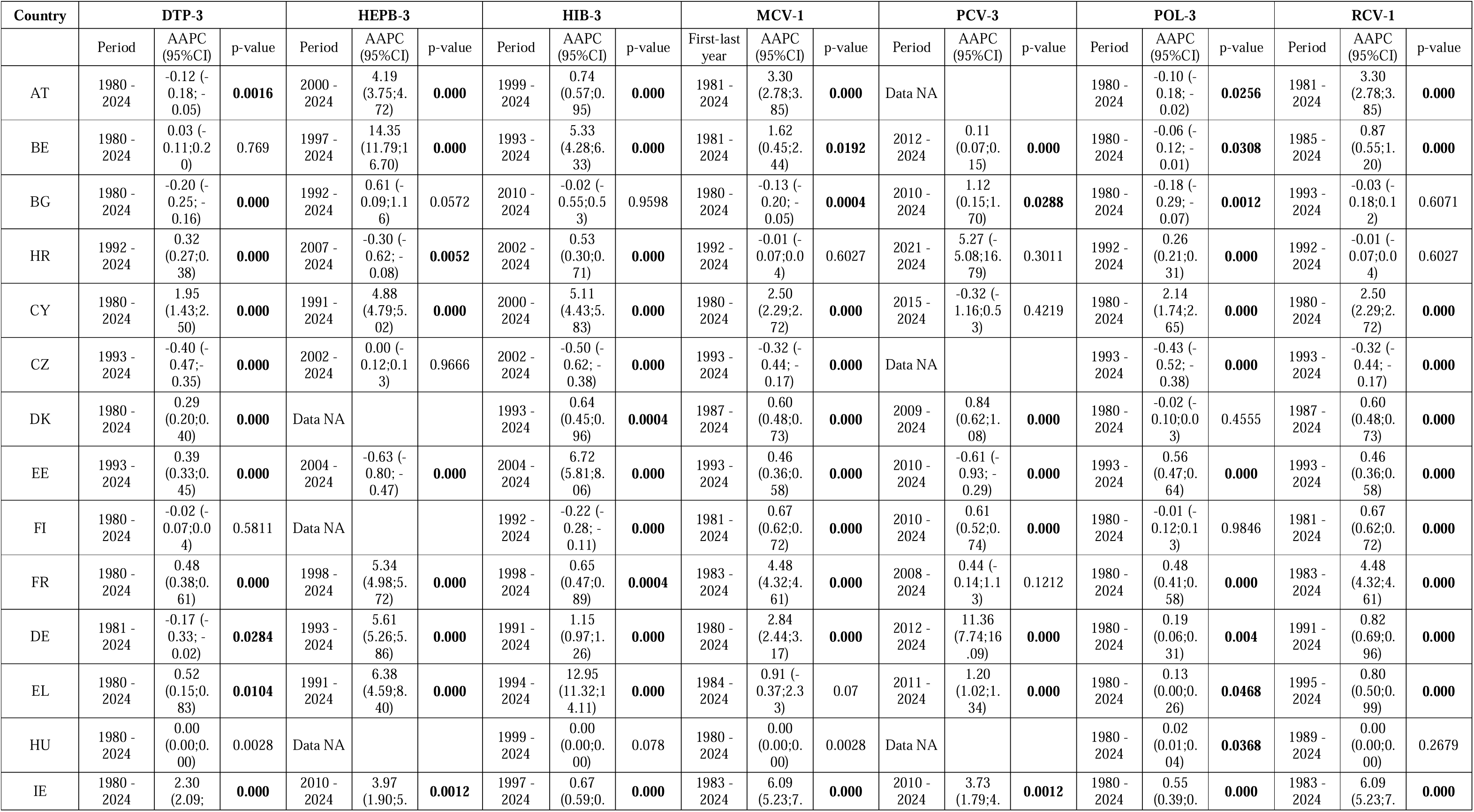
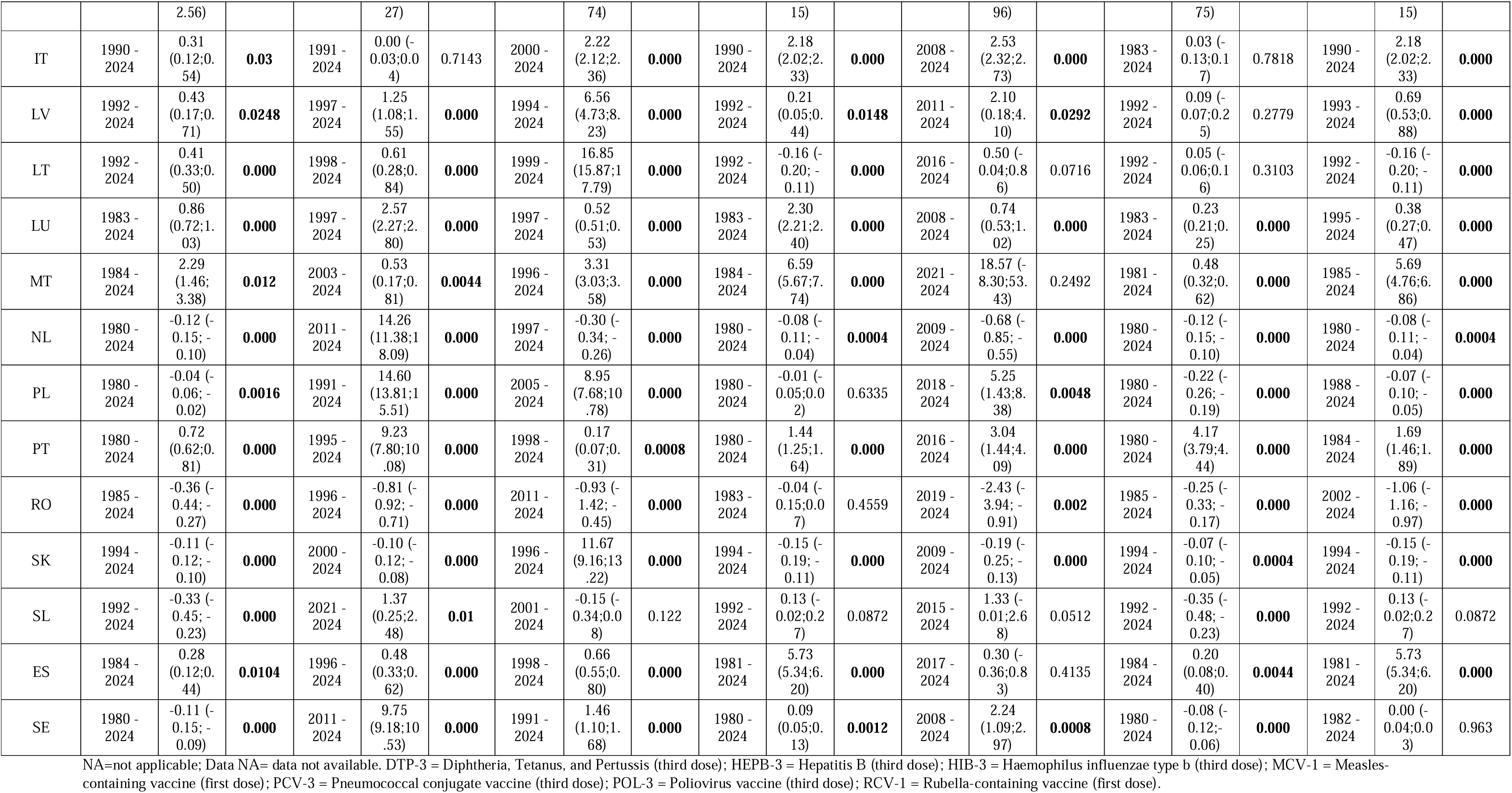
AAPC by antigen and country.

In the most recent year (2024), 71 country–antigen combinations achieved coverage ≥95%, while 62 were between 90% and 95% (Supplementary Table 3). Only Luxembourg reached ≥95% coverage for all antigens, and Hungary for all those reported (excluding hepatitis B and pneumococcal vaccination). Belgium and Latvia reached ≥95% coverage for 6 of 7 antigens. On the other hand, Romania reported coverage <80% for all antigens, while Estonia showed coverage levels around 80%.

### AAPC results by country

Overall, 20 countries had multiple positive trends in the AAPC for 4 or more coverages (Table 1). Among these, nine countries showed positive AAPCs for all seven vaccination coverages, including Greece, Ireland, Italy, Latvia, France, Luxembourg, Malta, Portugal and Spain.

Moreover, four countries (Belgium, Cyprus, Germany, Hungary) had positive AAPC for six different coverages, while Denmark, Estonia and Lithuania showed positive AAPCs for five different coverages. Finally, four countries (Austria, Croatia, Slovenia and Sweden) showed positive AAPCs for four different coverages.

On the contrary, some countries showed negative trends in the AAPC for multiple coverages. Romania showed a negative trend for all 7 coverages. The Netherlands and Slovakia showed negative trends for 6 coverages. Bulgaria and Czechia both showed negative trends for 5 coverages while Finland and Poland presented negative AAPC for 4 coverages.

### AAPC by antigen

The analysis of the AAPC by antigen revealed heterogeneous trends (Table 2). For DTP-3, 14 of 27 countries showed a significant increase, while 10 exhibited a significant decrease. For HEPB-3, 17 of 24 countries showed a significant increase, and 4 showed a significant decrease. HIB-3 trends displayed a clear predominance of significant increases, observed in 20 of 27 countries, with 4 significant and 2 non-significant decreases. For MCV-1, 16 of 27 countries showed a significant increase, while 5 presented a significant decrease and 3 a non-significant decrease. PCV-3 showed mainly significant (13) or non-significant (6) increases among 24 available countries, with only 4 significant and 1 non-significant decrease. POL-3 trends demonstrated a more balanced distribution, with 12 significant increases and 10 significant decreases across 27 countries. Finally, RCV-1 trends showed mostly significant increases (16 of 27 countries).

**Table 2.** AAPC by antigen.

| Antigen | Significant increase | Non-Significant increase | Significant Decrease | Non-Significant Decrease | No variation | Total |
| --- | --- | --- | --- | --- | --- | --- |
| DTP-3 | 14 (51.85%) | 1 (3.70%) | 10 (37.04%) | 1 (3.70%) | 1 (3.70%) | 27 |
| HEPB-3 | 17 (70.83%) | 1 (4.17%) | 4 (16.67%) | 0 (0.00%) | 2 (8.33%) | 24 |
| HIB-3 | 20 (74.07%) | 0 (0.00%) | 4 (14.81%) | 2 (7.41%) | 1 (3.70%) | 27 |
| MCV-1 | 16 (59.26%) | 2 (7.41%) | 5 (18.52%) | 3 (11.11%) | 1 (3.70%) | 27 |
| PCV-3 | 13 (54.17%) | 6 (25.00%) | 4 (16.67%) | 1 (4.17%) | 0 (0.00%) | 24 |
| POL-3 | 12 (44.44%) | 3 (11.11%) | 10 (37.04%) | 2 (7.41%) | 0 (0.00%) | 27 |
| RCV-1 | 16 (59.26%) | 1 (3.70%) | 6 (22.22%) | 2 (7.41%) | 2 (7.41%) | 27 |

DTP-3 = Diphtheria, Tetanus, and Pertussis (third dose); HEPB-3 = Hepatitis B (third dose); HIB-3 = Haemophilus influenzae type b (third dose); MCV-1 = Measles-containing vaccine (first dose); PCV-3 = Pneumococcal conjugate vaccine (third dose); POL-3 = Poliovirus vaccine (third dose); RCV-1 = Rubella-containing vaccine (first dose).

### Variation in the APC of trend A and trend B

The results revealed in total 335 positive APC and 213 negative APC (Supplementary Table 4). The analysis of the latest trends reveals predominantly negative evolution (110 cases), compared to positive evolution (40 cases). In 14 cases changes occurred between non-significant trends. Table 3 reports all the details about changes among trend A and B. Briefly, in 91 cases, trends shifted from positive to negative, and in 19 cases a positive trend lost significance. 16 trends maintained a significant decrease pattern. On the other hand, 16 trends shifted from negative to positive and in 1 case a negative trend lost significance. 23 trends maintained a significant increase pattern. Considering trends without joinpoint, it is observed an increase in 11 cases and decrease in 7 cases. *Vaccination coverage trends by country*

**Table 3.** APC for trend A and trend B, by country and antigen.

| Country | Metric | DTP-3 | HEPB-3 | HIB-3 | MCV-1 | PCV-3 | POL-3 | RCV-1 |
| --- | --- | --- | --- | --- | --- | --- | --- | --- |
| AT | Trend A | **-3.74** | **1.33** | **-3.3** | **35.95** | Data NA | **-3.77** | **36** |
|  | Trend B | -0.27 | **-1.08** | 0.16 | **1.44** | Data NA | -0.27 | **1** |
|  | Final coverage | 85% | 85% | 85% | 90% |  | 85% | 90% |
| BE | Trend A | 5.92 | **380.87** | **120.89** | **12.4** | **0.32** | **0.49** | **1** |
|  | Trend B | 0.07 | **1.94** | 0.09 | 0.85 | 0 | -0.04 | NA |
|  | Final coverage | 97% | 97% | 97% | 96% | 94% | 98% | 96% |
| BG | Trend A | **-0.2** | **11.69** | -0.02 | **-0.13** | **13.41** | **-0.18** | **0** |
|  | Trend B | NA | -0.09 | NA | NA | **-0.8** | NA | **-0.35** |
|  | Final coverage | 94% | 94% | 94% | 94% | 91% | 94% | 94% |
| HR | Trend A | **0.5** | 1.37 | **9.34** | **0.49** | 5.27 | **0.46** | **0.49** |
|  | Trend B | **-0.31** | **-0.52** | **-0.31** | **-0.46** | NA | **-0.3** | **-0.46** |
|  | Final coverage | 92% | 91% | 92% | 90% | 89% | 92% | 90% |
| CY | Trend A | **50.27** | **1.56** | **18.71** | **44.52** | -0.32 | **48.97** | **45** |
|  | Trend B | 0.15 | **-0.18** | -0.02 | **0.23** | NA | 0.14 | **0.23** |
|  | Final coverage | 94% | 94% | 94% | 99% | 82% | 94% | 99% |
| CZ | Trend A | **-0.38** | **-0.49** | **-0.47** | -0.55 | Data NA | **-0.45** | -0.55 |
|  | Trend B | **-4.76** | **-4.38** | **-4.41** | **-3.75** | Data NA | **-4.46** | **-3.75** |
|  | Final coverage | 86% | 86% | 86% | 89% | Data NA | 86% | 89% |
| DK | Trend A | -1.55 | Data NA | **-0.85** | **-3.13** | **2.67** | 1.07 | **-3.13** |
|  | Trend B | **0.58** | Data NA | **0.59** | **0.79** | **0.38** | 0 | **1** |
|  | Final coverage | 96% | Data NA | 96% | 94% | 96% | 96% | 94% |
| EE | Trend A | **-0.36** | **-0.27** | **97.93** | -0.23 | Data NA | **-0.28** | -0.23 |
|  | Trend B | **-3.77** | **-2.29** | -0.36 | **-1.13** | Data NA | **-2.96** | **-1.13** |
|  | Final coverage | 81% | 81% | 81% | 83% | 87% | 81% | 83% |
| FI | Trend A | **-2.97** | Data NA | **-3.32** | **0.19** | **-0.61** | -2.93 | **0.19** |
|  | Trend B | 0.17 | Data NA | 0.19 | **-0.3** | NA | 0.16 | **-0.3** |
|  | Final coverage | 91% | Data NA | 91% | 94% | 96% | 91% | 94% |
| FR | Trend A | **4.82** | **11.47** | **2.62** | **2.02** | 0.08 | **-2.24** | **2** |
|  | Trend B | -0.04 | **1.34** | -0.06 | **0.47** | **0.76** | **0.37** | **0.47** |
|  | Final coverage | 96% | 96% | 96% | 95% | 75% | 96% | 95% |
| DE | Trend A | **4.97** | **21.48** | **2.21** | **5.19** | **11.4** | **8.11** | **1** |
|  | Trend B | -0.24 | 0.08 | **-0.24** | 0.22 | **-1.03** | **-0.26** | -0.04 |
|  | Final coverage | 89% | 85% | 88% | 96% | 90% | 88% | 96% |
| EL | Trend A | **12.68** | **146.64** | **21.03** | **1.9** | **87.54** | **0.13** | **1** |
|  | Trend B | 0.3 | 0.22 | 0.82 | -0.43 | 0.34 | NA | **-0.53** |
|  | Final coverage | 95% | 92% | 99% | 91% | 99% | 95% | 91% |
| HU | Trend A | **0** | Data NA | 0 | **0** | 1.84 | **0.15** | 0 |
|  | Trend B | NA | Data NA | NA | NA | 0.02 | 0 | NA |
|  | Final coverage | 99% | Data NA | 99% | 99% | 99% | 99% | 99% |
| IE | Trend A | **2.09** | **37.15** | **0.69** | **105.12** | **40.28** | 1.42 | **105** |
|  | Trend B | -0.07 | -0.72 | **-0.33** | **0.71** | **-1.36** | -0.22 | **1** |
|  | Final coverage | 92% | 92% | 92% | 90% | 84% | 92% | 90% |
| IT | Trend A | **5.47** | **-0.91** | **4.06** | **10** | **0.87** | **5.9** | **10** |
|  | Trend B | 0 | 0.13 | **-0.11** | **0.45** | -0.23 | **-0.15** | **0.45** |
|  | Final coverage | 94% | 94% | 95% | 95% | 90% | 94% | 95% |
| LV | Trend A | **4.32** | **365.79** | **71.33** | **-0.37** | **2.1** | 0.09 | **6** |
|  | Trend B | 0.03 | 0.39 | 1.08 | **0.43** | NA | NA | -0.02 |
|  | Final coverage | 97% | 99% | 97% | 98% | 92% | 97% | 98% |
| LT | Trend A | **0.35** | **12.65** | **10.23** | 0.2 | -0.28 | **0.52** | 0.2 |
|  | Trend B | **-0.42** | **-0.34** | -0.62 | **-1.35** | **2.9** | **-0.41** | **-1** |
|  | Final coverage | 89% | 87% | 89% | 86% | 87% | 89% | 86% |
| LU | Trend A | **0.88** | -0.8 | **0.39** | **2.43** | 3.6 | **1.01** | 0.43 |
|  | Trend B | 0.02 | 0.32 | 0 | **0.3** | 0.17 | -0.01 | 0.01 |
|  | Final coverage | 99% | 98% | 99% | 99% | 97% | 99% | 99% |
| MT | Trend A | **37.42** | **5.17** | **11.02** | **101.67** | 18.57 | **9.56** | **138** |
|  | Trend B | **0.71** | 0.53 | -0.17 | **1.22** | NA | -0.09 | **1** |
|  | Final coverage | 97% | 97% | 97% | 88% | 97% | 97% | 88% |
| NL | Trend A | **0.53** | **79.47** | 0.09 | **-0.38** | **-0.38** | **0.53** | **-0.38** |
|  | Trend B | **-1.38** | -0.21 | **-1.87** | **-1.34** | **-1.85** | **-1.38** | **-1.34** |
|  | Final coverage | 91% | 89% | 89% | 89% | 88% | 91% | 89% |
| PL | Trend A | **-0.78** | **11.23** | **124.01** | **-0.91** | **19.22** | **-1.75** | **-0.91** |
|  | Trend B | 0.09 | **-0.53** | 0.09 | 0.39 | -1.11 | **1.05** | 0 |
|  | Final coverage | 94% | 87% | 94% | 92% | 89% | 86% | 92% |
| PT | Trend A | **3.87** | **6.85** | **3.5** | 6.67 | **14.44** | **64.02** | 6 |
|  | Trend B | **0.16** | 0.14 | **0.16** | 0.2 | -0.5 | **0.25** | 0.2 |
|  | Final coverage | 99% | 99% | 99% | 99% | 98% | 99% | 99% |
| RO | Trend A | **0.29** | **-0.72** | **-0.93** | **0.98** | **-2.43** | **0.55** | **1** |
|  | Trend B | **-0.97** | **-2.72** | NA | **-0.92** | NA | **-0.93** | **-3** |
|  | Final coverage | 79% | 79% | 79% | 78% | 78% | 79% | 78% |
| SK | Trend A | **0.22** | **0.22** | **16.77** | 0.03 | **-0.97** | **0.23** | 0.03 |
|  | Trend B | **-0.47** | **-0.47** | -0.16 | **-0.31** | 0.02 | **-0.47** | **-0.31** |
|  | Final coverage | 96% | 96% | 96% | 94% | 96% | 96% | 94% |
| SL | Trend A | **0.87** | **1.37** | **0.75** | 0.13 | **6.54** | **0.46** | 0.13 |
|  | Trend B | **-0.56** | NA | **-0.55** | NA | **-4.83** | **-0.62** | NA |
|  | Final coverage | 90% | 90% | 90% | 95% | 56% | 90% | 95% |
| ES | Trend A | **0.74** | **6.83** | **6.76** | **21.6** | **2.63** | **1.36** | **22** |
|  | Trend B | -0.17 | **-0.2** | **-0.11** | **0.27** | -0.62 | **-0.2** | **0.27** |
|  | Final coverage | 94% | 94% | 94% | 97% | 92% | 94% | 97% |
| SE | Trend A | **-0.04** | **22.49** | **29.99** | **0.13** | **23.87** | **-0.07** | **0.13** |
|  | Trend B | **-0.66** | -0.11 | **-0.15** | **-1.1** | -0.52 | **-0.9** | **-1.1** |
|  | Final coverage | 96% | 95% | 95% | 93% | 94% | 95% | 93% |

NA=not applicable; Data NA= data not available. DTP-3 = Diphtheria, Tetanus, and Pertussis (third dose); HEPB-3 = Hepatitis B (third dose); HIB-3 = Haemophilus influenzae type b (third dose); MCV-1 = Measles-containing vaccine (first dose); PCV-3 = Pneumococcal conjugate vaccine (third dose); POL-3 = Poliovirus vaccine (third dose); RCV-1 = Rubella-containing vaccine (first dose).

Considering the changes between trends A and B, 17 EU countries showed predominantly negative patterns, with four or more decreasing vaccination coverages (Table 4). The most severe worsening in vaccination trends was observed in Bulgaria, Germany, Greece, The Netherlands, Romania, and Sweden, each showing seven decreasing trends, while Croatia, Czechia, Estonia, Lithuania, Slovakia, and Spain each recorded six decreases.

**Table 4.**
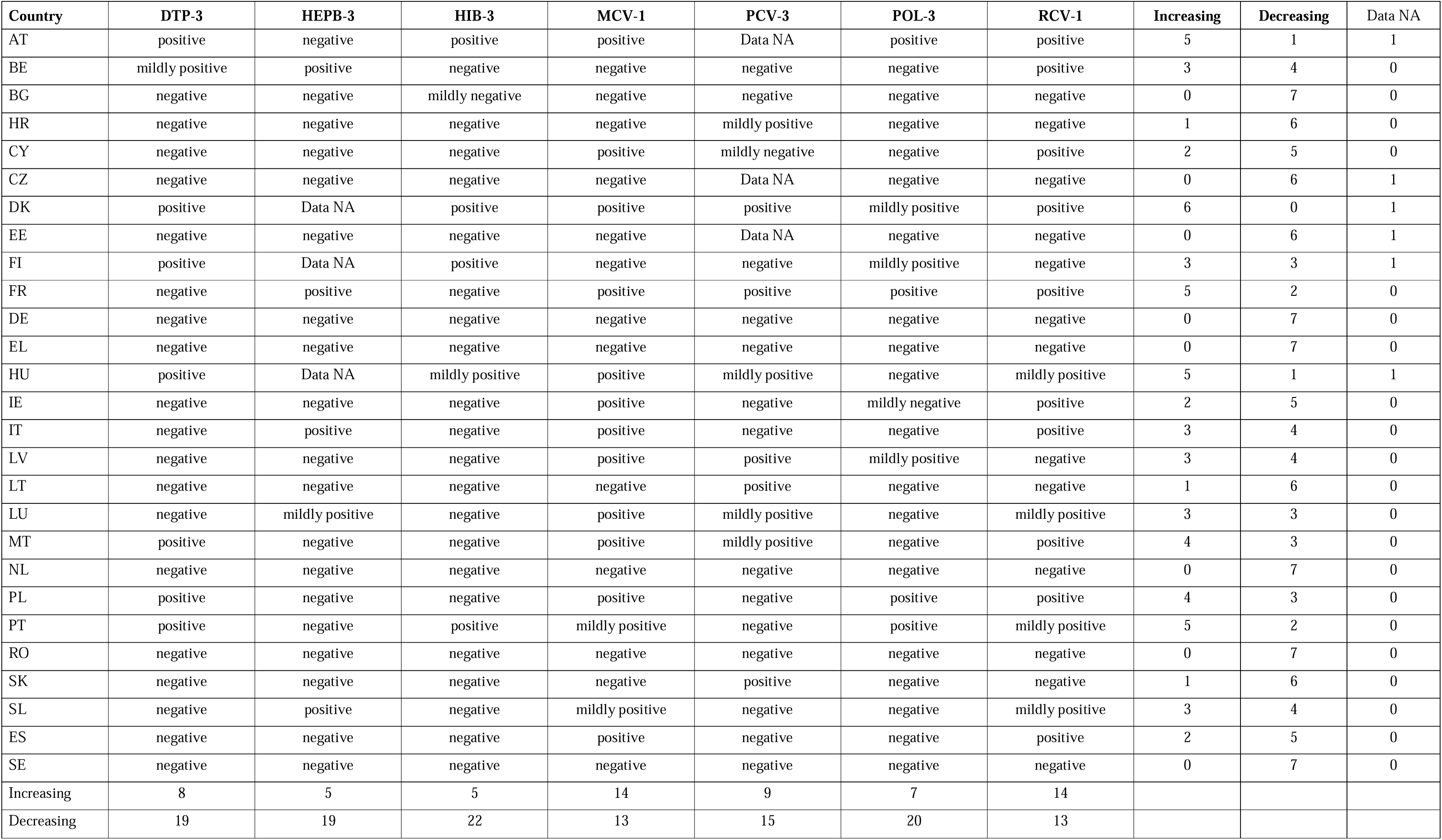
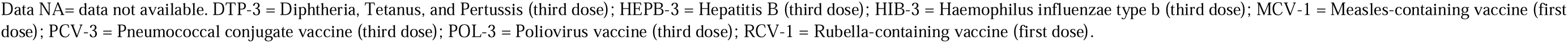
Increasing and decreasing trends considering changes from trend A and trend B, by country and antigen.

| Country | DTP-3 | HEPB-3 | HIB-3 | MCV-1 | PCV-3 | POL-3 | RCV-1 | Increasing | Decreasing | Data NA |
| --- | --- | --- | --- | --- | --- | --- | --- | --- | --- | --- |
| AT | positive | negative | positive | positive | Data NA | positive | positive | 5 | 1 | 1 |
| BE | mildly positive | positive | negative | negative | negative | negative | positive | 3 | 4 | 0 |
| BG | negative | negative | mildly negative | negative | negative | negative | negative | 0 | 7 | 0 |
| HR | negative | negative | negative | negative | mildly positive | negative | negative | 1 | 6 | 0 |
| CY | negative | negative | negative | positive | mildly negative | negative | positive | 2 | 5 | 0 |
| CZ | negative | negative | negative | negative | Data NA | negative | negative | 0 | 6 | 1 |
| DK | positive | Data NA | positive | positive | positive | mildly positive | positive | 6 | 0 | 1 |
| EE | negative | negative | negative | negative | Data NA | negative | negative | 0 | 6 | 1 |
| FI | positive | Data NA | positive | negative | negative | mildly positive | negative | 3 | 3 | 1 |
| FR | negative | positive | negative | positive | positive | positive | positive | 5 | 2 | 0 |
| DE | negative | negative | negative | negative | negative | negative | negative | 0 | 7 | 0 |
| EL | negative | negative | negative | negative | negative | negative | negative | 0 | 7 | 0 |
| HU | positive | Data NA | mildly positive | positive | mildly positive | negative | mildly positive | 5 | 1 | 1 |
| IE | negative | negative | negative | positive | negative | mildly negative | positive | 2 | 5 | 0 |
| IT | negative | positive | negative | positive | negative | negative | positive | 3 | 4 | 0 |
| LV | negative | negative | negative | positive | positive | mildly positive | negative | 3 | 4 | 0 |
| LT | negative | negative | negative | negative | positive | negative | negative | 1 | 6 | 0 |
| LU | negative | mildly positive | negative | positive | mildly positive | negative | mildly positive | 3 | 3 | 0 |
| MT | positive | negative | negative | positive | mildly positive | negative | positive | 4 | 3 | 0 |
| NL | negative | negative | negative | negative | negative | negative | negative | 0 | 7 | 0 |
| PL | positive | negative | negative | positive | negative | positive | positive | 4 | 3 | 0 |
| PT | positive | negative | positive | mildly positive | negative | positive | mildly positive | 5 | 2 | 0 |
| RO | negative | negative | negative | negative | negative | negative | negative | 0 | 7 | 0 |
| SK | negative | negative | negative | negative | positive | negative | negative | 1 | 6 | 0 |
| SL | negative | positive | negative | mildly positive | negative | negative | mildly positive | 3 | 4 | 0 |
| ES | negative | negative | negative | positive | negative | negative | positive | 2 | 5 | 0 |
| SE | negative | negative | negative | negative | negative | negative | negative | 0 | 7 | 0 |
| Increasing | 8 | 5 | 5 | 14 | 9 | 7 | 14 |  |  |  |
| Decreasing | 19 | 19 | 22 | 13 | 15 | 20 | 13 |  |  |  |

Data NA= data not available. DTP-3 = Diphtheria, Tetanus, and Pertussis (third dose); HEPB-3 = Hepatitis B (third dose); HIB-3 = Haemophilus influenzae type b (third dose); MCV-1 = Measles-containing vaccine (first dose); PCV-3 = Pneumococcal conjugate vaccine (third dose); POL-3 = Poliovirus vaccine (third dose); RCV-1 = Rubella-containing vaccine (first dose).

Nine countries exhibited moderately negative patterns, with four to five decreasing vaccines: Belgium, Ireland, Italy, Latvia, Poland, and Slovenia (four decreases each), and Cyprus, Finland, and Portugal (five decreases each).

In contrast, Denmark emerged as the best performer, with six increasing trends, followed by Hungary (five increasing trends and one decreasing), Austria and France, each showing five and four increasing trends, respectively. Malta also displayed a relatively balanced pattern, with four increasing and three decreasing trends. Finland and Luxembourg each had three positive and three negative trends, while Portugal presented five increasing and two decreasing coverages.

### Vaccination coverage trends by antigen

Considering the changes between trends A and B for each antigen, most vaccines showed predominantly decreasing patterns across EU countries, with MCV-1 and RCV-1 showing the most favorable evolution (Figure, 1, Figure 2, Table 4).

**Figure 1.**
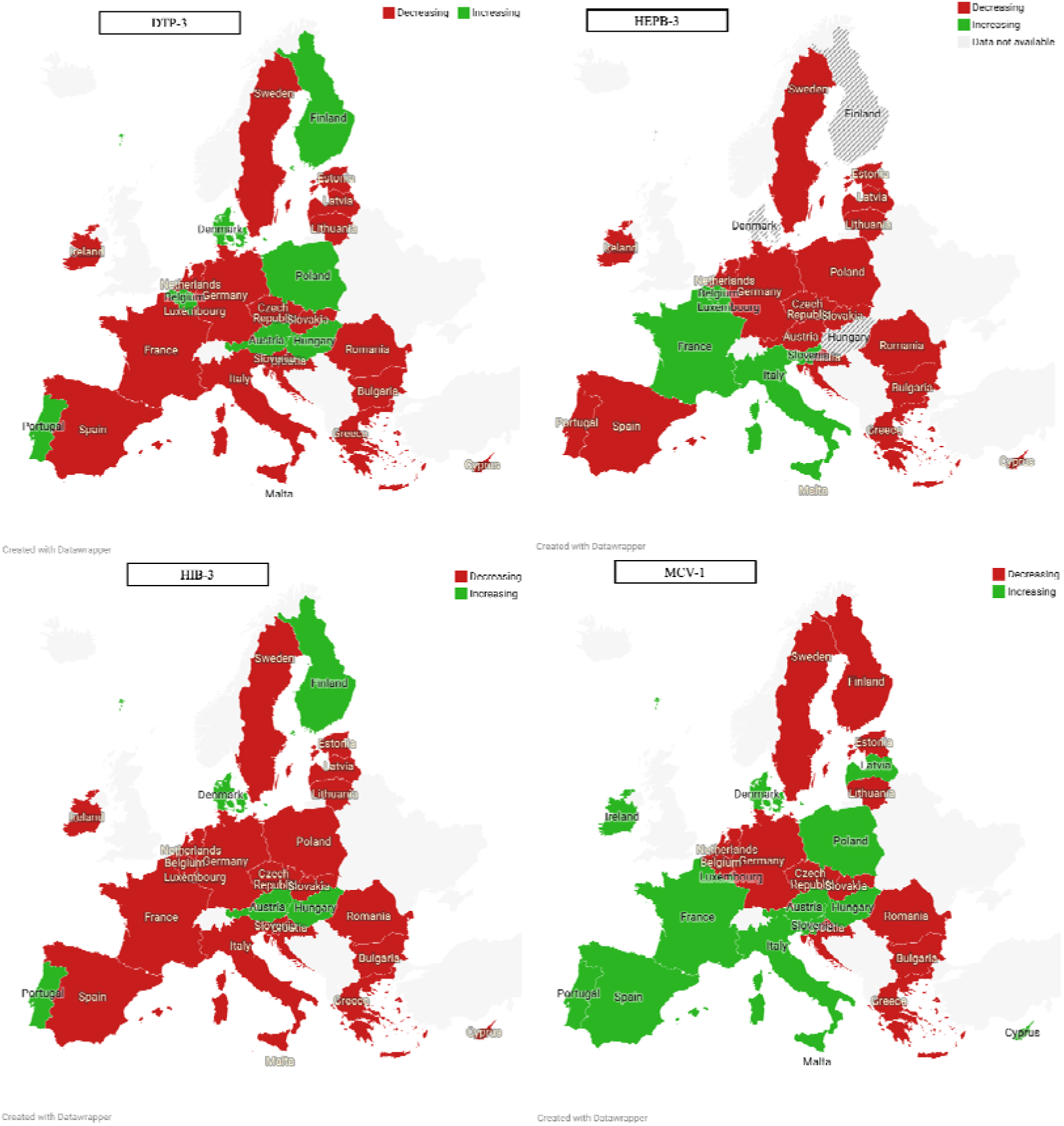
Variation in the APC of trend A and trend B in European Countries for DTP-3, HEPB-3, HIB-3, and POL-3.

**Figure 2.**
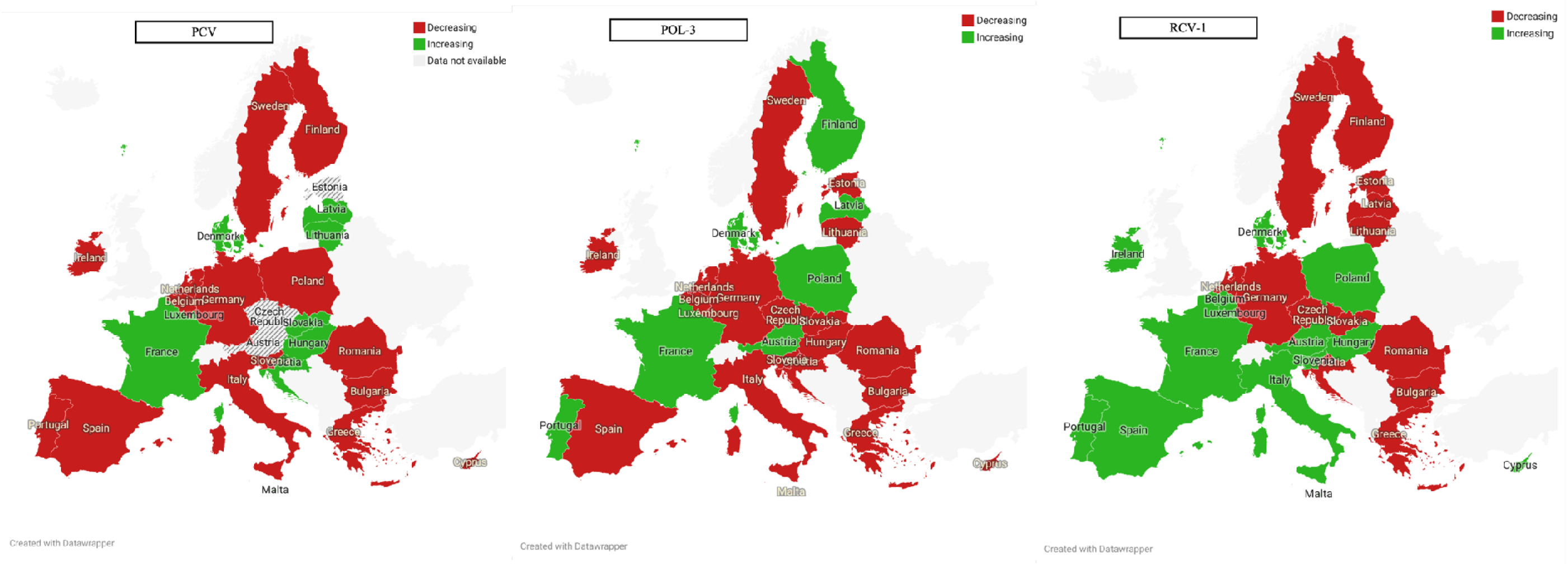
Variation in the APC of trend A and trend B in European Countries for PCV-3, MCV-1, and RCV-1.

The most marked decline was observed for HIB-3, with 22 countries showing decreasing trends, followed by POL-3 (20 countries), HEPB-3 (19 countries), and DTP-3 (19 countries). PCV-3 also exhibited a mainly decreasing pattern, with 15 countries showing declines, while MCV-1 and RCV-1 showed a more balanced situation, with 13 decreasing and 14 increasing trends each. Positive evolutions were less frequent overall. The highest number of increasing trends was observed for MCV-1 and RCV-1 (14 each), followed by PCV-3 (9), DTP-3 (8), POL-3 (7), HIB-3 (5), and HEPB-3 (5).

## Discussion

This study provides a comprehensive overview of vaccination coverage trends across the 27 EU Member States over more than four decades, highlighting the heterogeneity of temporal dynamics and recent reversals in long-term positive trajectories. Overall, while most countries have achieved and maintained high coverage for several antigens, recent years have been characterized by a slowdown or decline in vaccination rates. Our analysis revealed that although most countries displayed an increase over the full period, the most recent segmental analyses (trends A and B) indicated a predominance of negative evolutions, with only a few countries showing increasing trends, notably Denmark, Hungary, Austria, France, Malta, and Portugal. Moreover, heterogeneous patterns across vaccines were observed, with some (such as HIB-3, POL-3, and DTP-3) displaying widespread and alarming decreases. After decades of progressive improvement, vaccination coverage has recently stagnated or declined in a substantial number of EU countries. This finding aligns with previous reports from WHO and ECDC, which have documented a plateauing of routine immunization rates in Europe since the late 2010s, with the COVID-19 pandemic further amplifying the decline. The general decrease in vaccination coverage poses a substantial threat to public health (30), weakening herd immunity and exposing an increasing share of the population to vaccine-preventable diseases, particularly those at higher risk, such as infants, the elderly, and immunocompromised individuals (31,32). The potential consequences include the resurgence of controlled diseases and renewed outbreaks (31,33), leading to increased hospital admissions, higher healthcare costs, and additional strain on an already overstretched workforce (34,35).

The decline observed in recent years is likely multifactorial, reflecting a complex interplay of political, scientific, social, and cultural dynamics that have fuelled vaccine hesitancy, even in high-income settings (9,36). The rise of populist movements has further complicated this landscape, as political polarization has undermined trust in vaccination policies and institutions (37–39).

Moreover, inequities in access remain a persistent challenge. Disadvantaged populations are disproportionately affected by structural, geographic, and financial barriers to vaccination (40). Ensuring that vaccines are universally available and free of charge is therefore a fundamental equity measure, one that has consistently proven effective in improving coverage rates and is strongly recommended by WHO Europe.

Educational level is another well-documented determinant of vaccine acceptance (41–43), Addressing misinformation and knowledge gaps regarding vaccine safety and efficacy should be a policy priority. Introducing structured educational programs at both national and community levels, beginning in schools and involving children, parents, and teachers, can foster early awareness, promote health literacy, and build lasting confidence in immunization (44).

Making scientific evidence transparent, accessible, and understandable to the general population is also a crucial responsibility for public health professionals (45,46). As recommended by WHO Europe (47), insights into the barriers and enablers of vaccination should be translated into targeted, evidence-based interventions aimed at improving vaccine uptake. Healthcare professionals play a pivotal role in this process, as their knowledge, attitudes, and confidence are among the strongest determinants of their patients’ willingness to vaccinate (48,49).

Geographical patterns in vaccination coverage within the EU reflect differences in vaccination policies and practices, including the role of mandatory vaccination. Notably, six of the eight countries displaying predominantly positive trends have implemented some form of compulsory vaccination, an approach whose effectiveness is well documented (10,50,51). In other nations, however, such as France, where mandates were introduced only recently, or Germany and Greece, where mandates cover a limited number of vaccines, the full impact remains to be seen (52,53). While mandatory vaccination appears to be an effective countermeasure to declining coverage, these results should be interpreted with caution, given the influence of contextual, cultural, and political factors that are difficult to disentangle.

As widely recognized, prevention remains the only truly effective strategy to contain highly contagious diseases (54,55). Strengthening vaccination coverage must therefore be regarded as a public health priority. However, timely corrective action depends on the presence of active monitoring and robust surveillance systems capable of detecting and interpreting early warning signals (56). In this regard, the continuous analysis of vaccination coverage trends represents an essential component of infectious disease surveillance, providing the evidence base needed to guide policymakers, identify areas of concern, and investigate underlying causes (17).

### Limitations

This study has some limitations that should be acknowledged. First, we considered only the last two trends identified by the joinpoint model to classify vaccination trajectories and countries. While this approach may overlook earlier variations in long-term patterns, we believe that the most recent segments provide the most meaningful representation of the current situation and policy-relevant dynamics. Second, the Joinpoint regression relies solely on aggregated annual vaccination coverage data at the population level. As such, it does not allow for the identification of underlying causes or contextual factors driving observed changes in trends. Consequently, the associations between observed changes and specific explanatory variables could not be assessed. Third, our analysis focused primarily on temporal trends rather than absolute coverage levels. This means that countries such as Greece and Sweden, which displayed predominantly negative trends, still maintained high coverage levels, around or more than 90%. Nonetheless, even within countries with high baseline coverage, declining trends warrant careful attention, as they may indicate emerging vulnerabilities. Finally, countries maintaining coverage ≥95% and showing overall positive evolutions further validate the consistency and robustness of our analytical approach, confirming that trend-based analyses can complement absolute coverage indicators in identifying areas requiring public health attention.

## Conclusion

In recent years, vaccination coverage in most EU countries has declined, reversing the positive trend seen over the past forty years. This evidence calls for systemic countermeasures at international, national, and community levels, requiring a huge effort in which public health must play a leading role by providing scientific support to policymakers, communicating effectively, ensuring adequate education, and increasing the knowledge and skills of healthcare professionals on vaccines.

## Data Availability

All data produced are available online at https://www.who.int/teams/immunization-vaccines-and-biologicals/immunization-analysis-and-insights/global-monitoring/immunization-coverage/who-unicef-estimates-of-national-immunization-coverage

## Declarations

### Ethics approval and consent to participate

The study does not constitute human subjects research, nor does it use personal data of any kind. No Institutional Review Board approval was needed.

### Consent for publication

All Authors agree on the current version of the manuscript and give consent for publication.

### Availability of data and materials

All Correspondence and Material can be made available by the Corresponding Author upon reasonable request.

### Competing interests

The Authors declare no competing interests

### Funding

No funding was required for the study

## Acknowledgement of generative AI and AI-assisted technologies in the writing process

During the preparation of this work the author used a generative AI tool to facilitate the writing process. After using this tool, the authors reviewed and edited the content as needed and take full responsibility for the content of the publication.

## Author contributions

Conceptualization: LV; Methodology: LV, LR, FAC, CS; Software: LV, LR, CS, FAC; Validation: LV, WR, CDW, SB; Formal analysis: LR, FAC, CS; Data Curation: LV, LR, FAC, CS; Writing - Original Draft: LV, LR, FAC, CS; Writing - Review & Editing: LV, RP, WR, CDW, SB Visualization: LV; Supervision: LV, SB; Project administration: LV, SB.

## Notes

### Competing Interest Statement

The authors have declared no competing interest.

### Funding Statement

no funding received

### Author Declarations

WHO/UNICEF Estimates of National Immunization Coverage (WUENIC), https://www.who.int/teams/immunization-vaccines-and-biologicals/immunization-analysis-and-insights/global-monitoring/immunization-coverage/who-unicef-estimates-of-national-immunization-coverage

### Summary of Updates

The paper was updated with a thorough analysis including coverage data until 2024, according to latest availabilities.

